# Assessing Digital Health Maturity in Nigeria: Evidence from a Sub-national Assessment across ten (10) states and strategic imperatives for the health sector

**DOI:** 10.1101/2025.06.27.25330401

**Authors:** Chukwuemeka Azubuike, Chike Nwangwu, Ramat Seghosime, Precious Nwiko

## Abstract

Digital health technologies are pivotal for strengthening health systems and accelerating progress toward Universal Health Coverage (UHC). In Nigeria, while digital health interventions have been widely introduced, disparities in maturity and effectiveness are evident across different states. This study comprehensively assesses digital health maturity across ten Nigerian states, leveraging the Global Digital Health Index (GDHI) framework and the UN Foundation/ICT4SOML maturity model. Employing a mixed-methods approach that includes key informant interviews, document reviews, and quantitative scoring, the analysis evaluates seven core domains: leadership and governance, strategy and investment, legislation and compliance, standards and interoperability, infrastructure, workforce, and services and applications. The analysis also situates surveyed states along a maturity spectrum from experimentation to mainstreaming.

Findings reveal substantial variations in digital health readiness among the ten states. Lagos emerged as the most advanced, demonstrating robust governance structures, interoperability initiatives, and strategic investments in ICT infrastructure. In contrast, states like Gombe, Niger, Bauchi, Sokoto, Borno, Nasarawa and Yobe were the least matured, hindered by weak workforce capacity, poor infrastructure, and a lack of standardized interoperability. The assessment further identifies heavy reliance on donor-driven projects, limited state ownership, and fragmented governance as key barriers to sustainable digital health implementation.

To address these gaps, the study recommends establishing a National Digital Health Observatory for benchmarking and peer learning, alongside targeted state-level capacity building and infrastructure investments. The findings highlight the urgent need for stronger political commitment, structured governance, and the domestication of national digital health policies at the sub-national level. By mapping states on a digital health maturity spectrum, this study offers a clear framework for targeted interventions, optimized resource allocation, and accelerated progress toward a resilient, interoperable digital health ecosystem across Nigeria’s states.

## 1.0 Introduction

Global advancements in digital health have enabled healthcare delivery systems that deliver increased care accessibility, enhanced operational efficiency, and better health outcomes. The Global Strategy on Digital Health 2020–2025 highlights the importance of accessible digital solutions for building strong health systems. The strategy emphasizes the integration of digital technologies to enhance service delivery, reinforce governance structures, foster global collaboration, and accelerate progress toward universal health coverage.^1^

Integrating digital health interventions into national health systems is becoming increasingly important as health systems adapt to changing healthcare needs. In low- and middle-income countries (LMICs), including Nigeria, digital health technologies are increasingly considered critical to health system strengthening^2^. Since 2014, Nigeria has implemented the National Health Information and Communication Technology (Health ICT) Strategic Framework to reach Universal Health Coverage (UHC) by 2020 in the public sector^3^. The framework supports the combination of multiple health information systems, such as Civil Registration and Vital Statistics (CRVS)^4^ and National Health Management Information System (NHMIS), to enhance service delivery^5^.

Despite these efforts, Nigeria’s digital health ecosystem faces several challenges. Issues such as inadequate foundational infrastructure—e.g., digital identity, cybersecurity—internet connectivity problems, and a lack of standardized data interoperability and management practices persist, hindering the full potential of digital health solutions^6,7^. While health system stakeholders are optimistic about the potential of digital health to improve service delivery and accessibility, barriers such as low digital literacy and insufficient training in digital tools continue to impede progress^8,7^. These issues are more pervasive at the state level, where many digital health resources are being implemented but lack traction and may prove ineffective.

The COVID-19 pandemic highlighted these deficiencies, reinforcing the need for digitalized health services, particularly for surveillance, data monitoring, service delivery, and improved access to care. In the post-pandemic period, the private health sector in Nigeria has seen multiple startups and organizations emerge to support public initiatives through innovative digital health solutions, which include telemedicine, e-prescriptions, electronic medical records (EMR), inventory management, and laboratory management systems. The private sector plays an essential role in addressing longstanding deficiencies within the healthcare system by offering innovative, affordable, and accessible healthcare solutions that complement public sector efforts^9^,

A digital health landscape study by the UN Foundation showed that Nigeria is in the early adoption phase of digital maturity and would benefit from improved infrastructure readiness^10^. To chart a new digital health course for the country, the National Digital Health Strategy (NDHS) 2021-2025^11^ was developed by the Federal Ministry of Health in collaboration with digital health stakeholders. This strategy builds on the National Health ICT Strategic Framework. It sets out a practical roadmap to enable Health Information Exchange (HIE), guide policy and plans towards improving Nigeria’s digital health maturity.

So far, the varying implementation of the National strategy at the subnational level has led to fragmented efforts that make it difficult to establish a cohesive and sustainable digital health framework for adoption. This may be due to different digital maturity realities at the national and subnational levels. Overcoming these obstacles is essential for Nigeria to fully harness the potential of digital health technologies and improve healthcare outcomes for its population.

This paper examines digital health maturity at the sub-national level in Nigeria through a standardized framework that analyses digital health readiness and enabling environment across ten states. A similar sub-national digital maturity assessment has also been conducted in Kenya and was instrumental in identifying regional disparities in digital health infrastructure and capabilities, enabling county governments to tailor strategies that address specific local needs^12^. The aim of this study is thus to reveal the systemic and contextual barriers that prevent digital health interventions from being effectively implemented, especially at the state level, where numerous initiatives are tested but rarely continue. By mapping states along a maturity spectrum, the analysis reveals disparities and outlines practical pathways to accelerate progress towards digital health maturity in Nigeria.

## 2.0 METHODS

### Overview

The study employed a qualitative methodology that used key informant interviews and document reviews to gather insights from key stakeholders across states. This was complemented with a quantitative scoring system drawn from the Global Digital Health Index (GDHI) and World Health Organization-International Telecommunications Union (WHO-ITU) e-health strategy toolkit^13^, which allowed for a consistent way to compare progress across different domains.

### Conceptual Framework for Assessment

The assessment was underpinned by a conceptual framework that situated states along a five-stage maturity spectrum: experimentation, early adoption, development and building up, scale-up, and mainstreaming. This framework was adapted from the UN Foundation and ICT4SOML’s 2014 review, Assessing the Enabling Environment for ICTs for Health in Nigeria.

**Figure 1:**
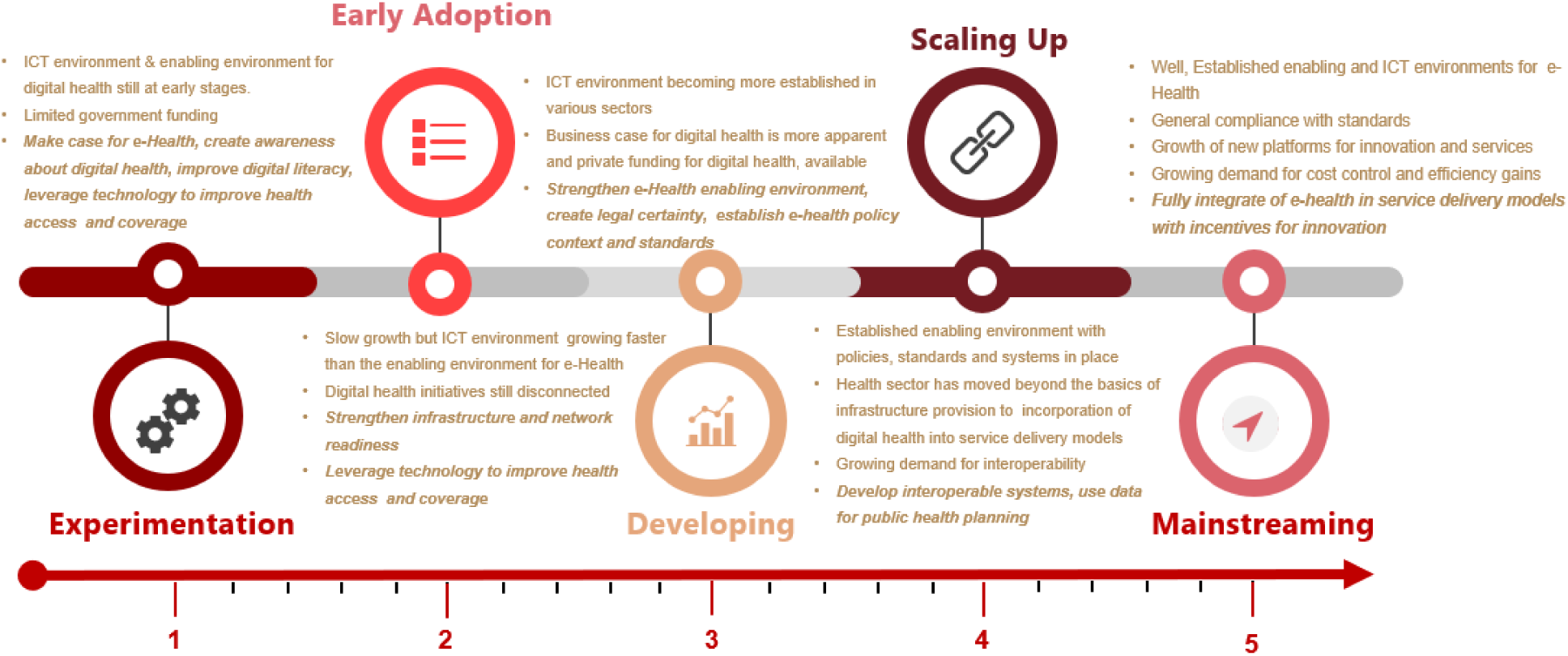
Maturity Spectrum adapted from UN foundation’s/ICT4SOML “Assessing the enabling environment for ICTs for health in Nigeria: a review of policies

The Global Digital Health Index (GDHI)/WHO-ITU framework was applied to determine each state’s position along this spectrum. This framework assesses digital health ecosystems across seven core domains: leadership and governance; strategy and investment; legislation and compliance; standards and interoperability; infrastructure; workforce; and services and applications. The first four domains assess the enabling environment, while the last three assess the ICT environment. Together, these constitute the building blocks of a national eHealth environment that will allow the eHealth outcomes to be achieved.

**Figure 2:**
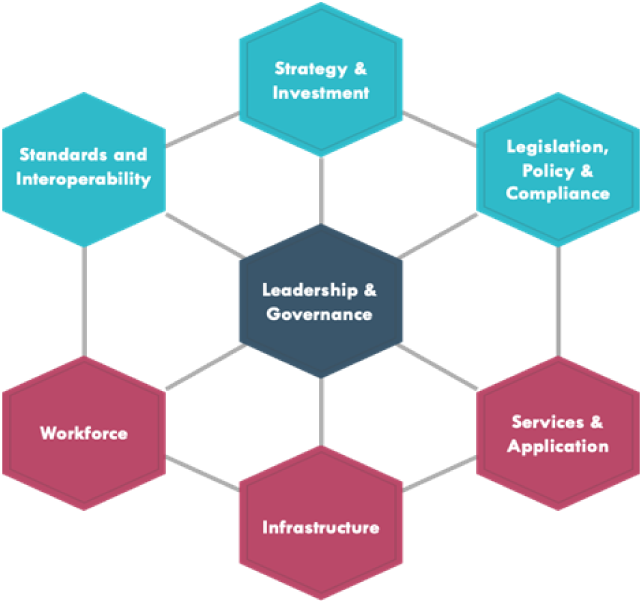
Global Digital Health Index/WHO-ITU framework depicting the components of a Digital health environment

### State Selection

The study was conducted in ten (10) states in Nigeria. These states consist of the focal program engagement states that have signed a memorandum of association (MOU) with the Gates Foundation in Nigeria and include Bauchi, Borno, Gombe, Kano, Kaduna, Lagos, Nasarawa, Niger, Sokoto and Yobe.

### Data Collection

Data was collected through key informant interviews with government officials, health professionals, and development partners and through desk reviews of existing digital health policies and programs. Responses were gathered using the Global Digital Health Index (GDHI) tool, covering 29 indicators across seven maturity domains.

#### Secondary data collection

A detailed review of the secondary data was conducted across the 10 states. Published and unpublished literature on digital health assessment and implementation was retrieved across the engagement states from 2015 to 2021^4^. The published materials were retrieved from databases such as Google Scholar, PubMed, Web of Science, etc. Relevant but unpublished studies were also sought actively from government stakeholders during the preliminary scoping meetings. Information from retrieved secondary data was collated and organized using an abstraction sheet that articulated digital health maturity and the 7 WHO/ITU domain areas. The abstraction template also organized information about entities’ structure, interaction, and roles in the health system and key barriers to widespread adoption and use of digital health at the national and state levels.

#### Primary data collection

Primary data was collected through key informant interviews (KIIs) at the national and subnational levels. These interviews aimed to capture firsthand perspectives on digital health implementation, system governance, and the operational environment across the ten focus states. A two-part assessment tool guided the data collection process.

Part A adapted the Global Digital Health Index (GDHI) to elicit structured responses across 19 indicators spanning the seven WHO–ITU digital health maturity domains: leadership and governance, strategy and investment, legislation and compliance, standards and interoperability, infrastructure, workforce, and services and applications. It collected responses to 19 indicators (and sub-indicators) across the seven key domain areas that constitute the WHO-ITU framework. These domain areas are building blocks or enablers whose effective functioning is necessary for digital health interventions’ success and long-term sustainability.

Part B gathered descriptive information on digital health innovations that respondents were directly involved in or familiar with at their respective levels. It used a semi-structured questionnaire to elicit the structure, interaction, and roles of entities in the health system and key barriers to widespread adoption and use of specific digital health at the national and state levels.

The interviews were conducted over a two-month period, between August-October 2021, by trained personnel and lasted 60-90 minutes per interview.

#### Respondent selection for primary data collection

Respondents for the primary data collection were selected from the ten focus states at the subnational level, using a purposive sampling approach. This approach ensured that individuals within decision-making positions, and with relevant experience and institutional knowledge in digital health, were captured in the study.

Respondents included ICT officers and advisors within State Ministries of Health and the offices of the respective state governors. Additionally, respondents were selected from state, zonal, and LGA-level monitoring and evaluation officers, as well as heads of drug management agencies (DMAs). A total of 18 stakeholders were interviewed at the national level, while approximately 270 stakeholders were engaged across the ten focus states.

**Table 1:** Description of respondents for the study at the state level.

| Category | Entity | Number of respondents |
| --- | --- | --- |
| **Health** |  |  |
|  | Department of Health Planning Research and Statistics | 1 |
|  | Department of Hospital services | 1 |
|  | Department of Public Health | 1 |
|  | National Primary Health Care development agency (NPHCDA) | 2 |
|  | National Agency for Food and Drugs Administration and Control (NAFDAC) | 1 |
|  | National Health Insurance Scheme (NHIS) | 2 |
|  | National Agency for the Control of AIDS(NACA) | 1 |
|  | National Institute of Medical Research (NIMR) |  |
|  | Centre for disease control (NCDC) | 1 |
|  | National Institute for Pharmaceutical Research and Development (NIPRD) |  |
|  | Office of the special assistant to the president on the MDGs |  |
|  | State Drug Management Agencies | 1 |
|  | Monitoring and Evaluation officers | 4 (1State, 1Zonal, 2LGAs) |
|  | Heads of health facilities at state level | 10 (5 public PHCs, 5 private hospitals) |
| **Sub-Total** |  | **25** |
| **ICT** | Department of e-government |  |
|  | National Information Technology Development Agency (NITDA) |  |
|  | Department of Planning Research and Statistics (DPRS) |  |
|  | National Communications Commission (NCC) |  |
|  | SA to the governor on ICT (where applicable) | 1 |
|  | State Ministry of ICT | 1 |
| **Sub-Total** |  | **2** |
| Total (National level + 1 state) |  | **27** |
| **Grand Total**<br>(Adjusted for 10 states) |  | **270** |

##### Interview Guide

| **PART A** |  |  |
| --- | --- | --- |
| **Domain** | **Indicator** | **Policy question** |
| **Leadership and governance** | Digital health prioritized at the state level through dedicated bodies / mechanisms for governance | Does the state have a separate department / agency / statel working group for digital health? |
|  | Digital Health prioritized at the state level through planning | Is digital health included and budgeted for in state health or relevant state strategies and/or plan(s)? The focus of this indicator is on the inclusion of digital health or eHealth in the state health strategy. |
| **Strategy and Investment** | State eHealth/ Digital Health Strategy or Framework | Does the country have an eHealth or digital health strategy or framework and a costed digital health plan? |
|  | Public funding for digital health | What is the estimated percent (%) of the annual public spending on health committed to digital health? |
| **Legislation, Policy and Compliance** | Legal Framework for Data Protection (Security) | Is there a law on data security (storage, transmission, use) that is relevant to digital health? |
|  | Laws or Regulations for privacy, confidentiality, and access to health information (Privacy) | Is there a law to protect individual privacy, governing ownership, access and sharing of individually identifiable digital health data? |
|  | Protocol for regulating or certifying devices and/or digital health services | Are there protocols, policies, frameworks or accepted processes governing the clinical and patient care use of connected medical devices and digital health services particularly in relation to safety, data integrity and quality of care? |
|  | Cross-border data security and sharing | Are there protocols, policies, frameworks or accepted processes in place to support secure cross-border data exchange and storage? This includes health-related data coming into a country, going out of a country, and/or being used in a country related to an individual from another country. |
| **Capacity building/Workforce** | Digital health integrated in health and related professional pre-service training (prior to deployment) | Is digital health part of curriculum for health and health-related support professionals in training, in general? |
|  | Digital health integrated in health and related professional in-service training (after deployment) | Specifically, is digital health part of curriculum for health and health-related support professionals in the workforce in general? [Defined as community health workers, nurses, doctors, allied health, health managers/administrators, and technologists] |
|  | Training of digital health workforce | In general, is training in digital health / health informatics / health information systems / biomedical informatics degree programs (in either public or private institutions) producing trained digital health workers? |
|  | Maturity of public sector digital health professional careers | Are there public sector professional titles and career paths in digital health? |
| **Standards and Interoperability** | State digital health architecture and/or health information exchange | Is there a national /state digital health (eHealth) architectural framework and/or health information exchange (HIE) established? |
|  | Health information standards | Are there digital health / health information standards for data exchange, transmission, messaging, security, privacy, and hardware? |
| **Infrastructure** | Network readiness | Extract the WEF Network Readiness Index score |
|  | Planning and support for ongoing digital health infrastructure maintenance | Is there an articulated plan for supporting digital health infrastructure (including equipment- computers/ tablets/ phones, supplies, software, devices, etc.) provision and maintenance? |
| **Services and Applications** | State scaled digital health systems | Are state scaled digital systems in place across specific priority programs/areas and are digital systems used to address them, and how many are at the national/state scale. |
|  | Digital identity management of service providers, administrators, and facilities for digital health, including location data for GIS mapping | Are health system registries of uniquely identifiable providers, administrators, and public facilities (and private if applicable) available, accessible, and current? Is the data geotagged to enable GIS mapping? |
|  | Digital identity management of individuals for health | Are secure registries or a master patient index of uniquely identifiable individuals available, accessible, and current for use for health-related purposes? |

PART B *Additional Questions about specific digital health projects identified by respondents*
|  | Section | Question |
| --- | --- | --- |
|  | Structure | Can you discuss the structures of these programs/interventions?<br>Who were the drivers of these programs?<br>What were the roles of the different entities involved in the implementation (State level MDAs, donors, and private sector organizations)?<br>What non-governmental organizations contributed to these implementations? |
|  | Communication | How would you describe the type of interactions that exist between the relevant TWGs, ministries, agencies, departments, and other organizations?<br>During the implementation of these interventions, how would you assess the relationship between the SMOH, SMICT and your Department/Agency?<br>How would you assess the ease of information sharing between SMOH, SMICT, and your agency/department?<br>What are the challenges that exist in communication? |
|  | Barriers to Adoption | • Briefly discuss some of the barriers to the adoption of these interventions in the State?<br>• What solutions were adopted?<br>• Was a proper risk analysis done before implementation? Were there stakeholders that were not carried along? What would we do differently? |
|  | Critical Success Factors | • What are some of the factors that contributed to the success of the interventions?<br>• What would you say are factors that will drive the success of a fully scaled digital health system at the National/State-wide level? |
|  | Lessons Learnt | Please discuss some of the lessons learnt from the implementation of these interventions in the State. |
|  | Recommendations | • What are your recommendations for the r State to achieve success in digital health implementation?<br>• Are there any other important issues that we should have discussed which were not brought up? |

#### Training and Fieldwork

Before data collection began, field enumerators underwent a three-day training course. The sessions were held virtually via the Zoom conferencing platform and lasted approximately six hours each day. To ensure coordination and engagement, enumerators were grouped into two cohorts. Group 1 comprised teams responsible for the first five focus states, while Group 2 covered the remaining five states.

The first two days of training focused on familiarizing the enumerators with the assessment tool, the rationale of the study, expected outputs, and ethical considerations in data collection. Enumerators were also guided on interview etiquette and strategies for engaging diverse stakeholders. On the third day, responsibilities were assigned, and final preparations were made for the fieldwork phase.

#### Data Quality Assurance

Data collection was conducted over 30 days across state-level respondent organizations. Four supervisors from the core project team were assigned oversight roles and provided continuous support to field teams throughout the data collection process. Data was collected electronically using mobile devices pre-loaded with the GDHI questionnaire via the Open Data Kit (ODK) app. Collected data was transmitted to the cloud, and submissions were pre-assessed in real time for completion. A control room of analysts was set up to analyze the data for inconsistencies.

#### Data Analysis

94 respondents participated in the assessment. Data from the Part A survey was exported and cleaned in Microsoft Excel. Responses were used to generate scores for each of the seven domains based on the GDHI framework, which were then averaged to produce an overall digital health maturity score for each state.

To minimize the effects of recall bias, greater weight was given to verifiable documentary evidence when inconsistencies arose between interview responses and secondary data. For example, if the respondent’s input suggested a lower domain score but supporting documents from the desk review indicated a higher score, the latter was prioritized.

After GDHI scoring, each state was situated along a five-stage spectrum from experimentation, early adoption, development and build-up, scale-up, and mainstreaming. The parameters for situating each state on the maturity spectrum were based on the benchmarks outlined in the UN Foundation/ ICT4SOML framework. Parameters included: critical components such as ICT infrastructure, enabling policies, digital literacy, and health information systems integration. This analytical approach involving the GDHI and the UN Foundation/ICT4 SOML frameworks allowed for a better understanding of where each state stands in its digital journey and the steps required to progress along the maturity spectrum.

In Lagos State, direct interviews with government stakeholders could not be conducted due to an ongoing digital health assessment led by the state. In this case, responses were triangulated from interviews with four donor and private sector partners actively implementing digital health projects in Lagos.

The scoring process was complemented by a thematic analysis of qualitative data collected through Part B of the tool. Interview transcripts were reviewed, coded, and analysed using NVivo 12. A codebook was developed to guide the analysis, focusing on key dimensions such as structure, coordination, barriers to adoption, success factors, lessons learned, and recommendations. The structured coding allowed for systematically exploring emerging patterns across states and stakeholder types.

#### Ethical Considerations

Ethical approval was obtained from the NationalHealth Research Ethics Committee at the Federal Ministry of Health (FMOH). All qualified field data collectors were trained per ethical standards and best practices relevant to the study. Informed written consent was obtained from all participants before data collection. Individuals who declined to give consent were excluded from the study. Participant confidentiality was maintained throughout the process, and all data were stored securely.

## 3.0 FINDINGS

### 3.1 Overview of Performance on the GDHI Index

The digital health maturity assessment evaluated the capacity and readiness of ten Nigerian states using the Global Digital Health Index (GDHI) framework, which covers seven core domains: leadership and governance, strategy and investment, legislation and compliance, standards and interoperability, infrastructure, workforce, and services and applications. Each domain was scored on a five-point scale, and the average of these scores was used to position each state along a five-stage maturity spectrum, ranging from experimentation to mainstreaming. Scoring was based on aggregated responses from key informants, triangulated with documentary evidence, and validated through a standardized assessment process.

As shown in Table 2 below, leadership & governance and services & applications were the most mature domains across the states. These domains benefited from the some adoption of national systems like DHIS2 and the presence of at least nominal leadership structures in some states. Even in lower-performing contexts, digital health was often recognized as a policy priority, though implementation mechanisms remained weak.

**Table 2:** Table showing comparative scores per domain and overall GDHI score per state.

| State | Leadership & Governance | Strategy & Investment | Legislation & Compliance | Standards & Interoperability | Infrastructure | Workforce | Services & Applications | Average Score (Across domains) | Maturity Stage |
| --- | --- | --- | --- | --- | --- | --- | --- | --- | --- |
| Lagos | 3 | 2 | 2 | 3 | 2 | 2 | 2 | 2.3 | Early Adoption |
| Kaduna | 2 | 1 | 2 | 1 | 2 | 2 | 2 | 1.7 | Experimentation |
| Kano | 2 | 2 | 1 | 1 | 1 | 1 | 3 | 1.6 | Experimentation |
| Bauchi | 1 | 1 | 1 | 1 | 1 | 1 | 2 | 1.1 | Experimentation |
| Borno | 1 | 1 | 1 | 1 | 1 | 1 | 2 | 1.1 | Experimentation |
| Gombe | 1 | 1 | 1 | 1 | 1 | 1 | 1 | 1.0 | Experimentation |
| Nasarawa | 1 | 1 | 1 | 2 | 1 | 1 | 1 | 1.0 | Experimentation |
| Niger | 1 | 1 | 1 | 1 | 1 | 1 | 1 | 1.0 | Experimentation |
| Sokoto | 1 | 1 | 1 | 1 | 1 | 1 | 1 | 1.0 | Experimentation |
| Yobe | 2 | 1 | 1 | 1 | 1 | 1 | 1 | 1.0 | Experimentation |
| **Domain Score (Average)** | **1.5** | **1.2** | **1.2** | **1.3** | **1.2** | **1.2** | **1.6** |  |  |

Conversely, **standards and interoperability**, **infrastructure**, and **workforce** domains showed consistently low performance across most states. These weaknesses include the lack of formal digital architecture, inadequate power and connectivity infrastructure, and structured training or career pathways for digital health professionals. For instance, many states lacked any articulated plan for maintaining digital systems, and most respondents were unaware of existing interoperability frameworks beyond DHIS-2. Similarly, digital health capacity-building efforts were largely donor-driven and fragmented, with no incorporation into pre-service training curricula.

Strategy, investment, and legislation domains scored poorly across states. While most states had some exposure to the National Digital Health Strategy or broader ICT initiatives, only a few had budgetary provisions specifically targeting digital health. Moreover, although national-level policies like the NDPR were acknowledged, awareness and enforcement at the state level were limited.

When comparing states, Lagos, Kaduna, and Kano demonstrated relatively strong performance across multiple domains, particularly in leadership, infrastructure, and services. These states had more active donor engagement, visible systems investment, and early efforts toward interoperability via the Smart Health Interoperability Platform (SHIP) in the case of Lagos. States such as Gombe, Niger, Sokoto, Nasarawa, and Yobe showed more limited progress, particularly in the foundational domains of infrastructure and workforce development.

### 3.2 Domain-Specific Analysis

#### 3.2.1 Leadership and Governance

The leadership and governance domain assesses the extent to which digital health is prioritized and institutionalized at the state level through dedicated coordinating bodies or technical working groups (TWGs). The assessment revealed that in most states, such structures are either non-existent or non-functional, limiting the capacity to drive and sustain digital health efforts.

Across the ten states assessed, few had any formal body or agency overseeing digital health implementation. In Borno state, a modest effort was made to form an eHealth working group under the Department of Planning, Research, and Statistics, with some support from partners such as eHealth Africa. However, coordination platforms such as the Health Data Consultative Committee and other ad hoc structures meet infrequently and lack a clear mandate or continuity. At the time of the study in 2021, the last known TWG meeting in Borno was held in 2018, reflecting a broader pattern of irregular engagement.

In states such as Bauchi, Nasarawa, Yobe, Gombe, and Sokoto, respondents consistently reported the absence of any state-level coordinating mechanism for digital health. In these contexts, digital health activities—where they exist—are often donor-driven and implemented without a clear link to state-owned plans or oversight structures. Leadership structures were not visible even in relatively better-performing states like Kaduna and Kano. In Kaduna, apart from the executive governor having a dedicated special advisor on ICT, there were no formal or documented governance structures for digital health. Respondents often referred to the Kaduna Bureau of Statistics (KDBS), ICT units, M&E directorates, or general health planning departments as the closest institutional home for digital health, but these lacked the dedicated capacity or authority to coordinate digital health across programs and were not the dedicated governance mechanism for digital health coordination.

Lagos State showed relatively stronger commitment, with stakeholders referencing structured efforts such as the Smart Health Information Platform (SHIP), budgetary lines for ICT infrastructure, and the existence of a state-led and strong technical working group for digital health/ICT in the state.

#### 3.2.2 Strategy and Investment

The strategy and investment domain assesses whether states have developed formal digital health strategies and costed action plans and allocated dedicated budget lines for implementation. It examines the balance between donor-led and government-led investments in digital health across states.

Across all ten states canvassed, there was no clear evidence of comprehensive, state-owned digital health strategies or costed action plans. Digital health was often not reflected in state health sector strategies or annual operational plans. States such as Gombe, Borno, Bauchi, Sokoto, and Nasarawa were noted to lack digital health-specific strategies or frameworks. Where references to digital health appeared in development plans (e.g., Borno’s 25-year Development Plan), documentation was either inaccessible or lacked specific implementation guidance.

Budgetary allocations for digital health were also absent. While some states included line items for ICT equipment, these were typically generic and unrelated to digital health programs. For instance, in Bauchi and Sokoto, allocations for the procurement of ICT equipment were present in the budget. Still, no evidence suggested they were directed toward digital health systems or applications. A few respondents in Kaduna and Lagos acknowledged budgetary support for digital infrastructure and digital technology.

Without state-led investment, donor organizations and development partners predominantly drove digital health activities. States such as Bauchi, Borno, and Gombe reported that most digital health interventions— including the provision of hardware, software, and capacity-building—were funded and implemented by partners such as eHealth Africa, USAID, BMGF, and UNICEF. In Gombe, for example, the Go-Health platform for health insurance enrollment was developed with funding support from the State and the Basic Healthcare Provision Fund (BHCPF). Still, broader scale-up of digital modules (e.g., for claims or purchasing) was constrained by a lack of funding.

States such as Kaduna and Lagos still heavily rely on donor support for core infrastructure and digital implementation. In Lagos, despite political interest and more defined budgeting for ICT in health, digital health remains largely financed through blended partnerships, with private sector actors supporting digital initiatives such as the SHIP platform.

#### 3.2.3 Legislation, Policy, and Compliance

This domain assesses the presence and enforcement of data protection and privacy laws at the state level, the extent of awareness and application of the National Data Protection Regulation (NDPR), and the alignment of state-level practices with national digital health policy frameworks.

There was a general low awareness of the NDPR across many states, particularly in the public sector. In many states, respondents reported no knowledge of the NDPR or a vague understanding of its role and importance. Private sector actors, however, demonstrated higher levels of awareness and compliance, with many referencing internal data protection standards and audit practices aligned with NDPR requirements. In Lagos, for instance, private sector stakeholders confirmed that they complied with data audits mandated by the National ICT Development Agency (NITDA). A respondent was quoted as saying *“NITDA has been pushing compliance with the regulations through the data audits across data organizations”*

Furthermore, formal data governance laws at the state level were largely absent. While some states mentioned generic data policies or confidentiality clauses within civil service codes, none had enacted state-specific legislation governing digital health data security, privacy, or cross-border sharing. Gombe, Nasarawa, Yobe, and Sokoto states had no identifiable policies for regulating data storage, transmission, or cross-border exchange. In a few cases, like Borno, there were isolated procedural norms driven by donors or implementing partners, such as WHO- or eHealth-supported platforms with built-in data standards, but these lacked formal legal backing. In Bauchi state, respondents stated that there were laws that govern data security within the state. However, these laws may not be specific to digital health; they may apply to the security of general data.

In terms of alignment with national digital health policy, most states had not domesticated national frameworks or policies such as the National Digital Health Strategy (2021–2025). Although some states, such as Kaduna and Lagos, demonstrated early-stage translation of national policies, this had yet to translate into clear legal or compliance frameworks within state systems. A few states, including Nasarawa and Borno, mentioned efforts to domesticate the NDPR, but progress remained limited and inconsistent.

#### 3.2.4 Standards and Interoperability

This domain assesses whether states have adopted digital health architectural frameworks, implemented protocols for system interoperability, and benefited from integrated platforms supported by government, private sector, or donor initiatives.

Awareness and adoption of interoperability frameworks were minimal across the states. Responses from the stakeholders revealed they had a poor understanding of a digital health architectural framework, and they often appeared to conflate the subject with HMIS systems. Many respondents, especially from state-level government agencies, were unaware of any digital health architecture, health information exchange, or guidelines for integrating systems. Only Lagos state made a notable reference to an interoperability initiative: the Smart Health Information Platform (SHIP), which is designed to serve as a unified data warehouse for all digital health applications in the state. The platform intends to interface with existing systems through application programming interfaces (APIs) and assign a unique SHIP ID to patients, linked to their national identity number (NIN) and bank verification number (BVN). While promising, the platform at the time of this study in 2021 was still in the conceptual or early implementation phase during the assessment period.

In Kaduna, platforms like the Health Facility Analytics (HEFA) and the data quality assessments dashboard were developed with donor support. They provided spatial and programmatic insights, but integration with other systems was poorly established. Similarly, Bauchi and Gombe referred to digital repositories and standalone tools, but these operated in silos without harmonized data exchange protocols. In Niger and Sokoto states, there was no mention or clear evidence to show the existence of health information standards. The District Health Information System (DHIS-2) emerged as the most widely used and recognized platform for health information management. However, many respondents were not aware of how DHIS-2 could enable interoperability.

The role of donors and private sector actors in supporting data platforms and integration was evident across several states. In Kaduna, Sokoto, and Borno, partners such as eHealth Africa, the Gates Foundation, and UNICEF were central to deploying tools like Electronic Infectious Disease Surveillance and Response (EIDSR) and other logistics or surveillance systems. However, these systems were typically developed to meet the needs of specific programs and lacked integration with broader state health information systems.

#### 3.2.5 Infrastructure

The infrastructure domain examines the physical and digital elements required to support digital health systems, including internet connectivity, power supply, cloud systems, availability of ICT hardware, and mechanisms for system maintenance and support.

Across the ten states assessed, digital health infrastructure was consistently weak, with most states lacking an articulated plan to establish and sustain their digital health infrastructure. Basic hardware, such as computers, tablets, and servers, was often unavailable or insufficient, especially at the local government and facility levels. In some of the surveyed states, such as Borno, Gombe, and Bauchi, equipment provision was largely concentrated in specific donor-supported programs and rarely extended to the broader health system.

Power supply emerged as a critical bottleneck in nearly all states surveyed. Health facilities, particularly rural areas, faced frequent or prolonged electricity outages, often relying on generators without a consistent fuel supply. This unreliability limited the use of digital tools that require constant power or regular recharging. Some states, such as Kaduna and Lagos, had facilities equipped with solar power, which offered partial solutions to power-related disruptions, but these were not widespread. Internet connectivity was also limited, especially outside urban centers. Many facilities operated in low-bandwidth environments, which delayed data entry and hindered real-time system functionality. In Niger, respondents attested to the presence of plans to provide digital health infrastructure, especially hardware but implementation was limited by costs involved.

Maintenance support for ICT equipment was inconsistent and heavily donor dependent. In most states, government entities lacked the technical capacity to maintain hardware, host, and troubleshoot software systems. Instead, donor partners are often responsible for initial deployment and limited support. Once external support concluded, systems often fell into disuse due to a lack of ongoing maintenance or replacements.

A few states made significant progress in implementing GIS and facility mapping systems. The SHIP and HEFA platforms operated in Lagos and Kaduna, respectively, included spatial mapping components. In Borno and Yobe, health facilities were being geo-tagged under initiatives supported by WHO, eHealth Africa, or other implementing partners. However, across several states, data on facility coordinates were either incomplete or outdated. Where mapping had occurred, the databases were often not publicly accessible or linked to other health information systems.

#### 3.2.6 Workforce

The workforce domain assesses the extent to which digital health competencies are embedded within the state health workforce through training, professional development, and clear institutional career pathways.

Across the surveyed states, there was no evidence of structured pre-service training in digital health for health professionals. Most respondents across states, including Kaduna, Nasarawa, Gombe, and Yobe, indicated that digital health is not yet included in the curriculum of training institutions. While isolated programs in a few institutions (e.g., Colleges of Health Technology in Borno and Kaduna) reportedly offered modules on medical records, these offerings were not specific to digital health.

In-service training was more common but generally donor-led and program-specific. Most training focused on tools like DHIS2 or program-based surveillance systems and was concentrated among M&E officers and data clerks. For example, USAID-supported capacity-building efforts were active in states like Kebbi, Sokoto, and Bauchi, targeting health information system workers. In Lagos, donor partners and private actors conducted workshops and mentorship programs, particularly for health workers involved in digital interventions. However, these trainings were limited in scope and did not form part of a structured or continuous capacity development plan.

There was a notable absence of career paths or professional titles for digital health roles within the public sector. None of the states assessed had formalized positions, such as health informaticians or digital health officers, or promotion structures that recognized expertise in digital health. Some respondents pointed to roles such as M&E officers, HMIS managers, or ICT officers as having responsibilities relevant to digital health. Still, these roles relate more to health data management than digital health. Even in relatively mature digital contexts like Lagos, the concept of digital health as a distinct specialization for public health workers had not been institutionalized.

Digital literacy among health workers was generally low, especially at the primary and secondary healthcare levels. Respondents across states noted that many health workers were uncomfortable using basic ICT tools, which negatively affected data entry quality and limited the uptake of digital systems. Even when trained, many frontline workers struggled to retain or apply knowledge, especially in environments lacking infrastructure or supervisory support.

#### 3.2.7 Services and Applications

This domain evaluates the existence and scope of digital health applications across states, including patient management systems, health registries, surveillance tools, and platforms supporting data use for service delivery.

All ten states had some exposure to nationally deployed platforms, most notably DHIS2, which remains the most widely implemented digital health tool across the country. In every state, DHIS2 was used—at least partially—for routine health data reporting, particularly at the state and LGA levels. However, implementation was often limited to vertical health programs. In many cases, data continued to be collected manually before being entered into the system, delaying its usefulness for real-time decision-making.

Beyond DHIS2, the presence of other digital applications varied significantly by state. Kaduna exhibited a more diversified portfolio, including the Health Facility Analytics (HEFA), Family Planning dashboard, DQA dashboard, and Integrated Supportive supervision (ISS) platform. These tools were developed with donor partners to visualize service delivery data, support planning, and monitor performance. Similarly, Lagos was home to several facility-level tools such as Helium Health EMR and eHealth applications that support patient management, billing, and operational workflows in some public and private health facilities. These were complemented by efforts to create a unified platform through the SHIP initiative.

In other states, digital health applications were generally limited to program-specific use cases. For example, Borno and Sokoto utilized the Electronic Integrated Disease Surveillance and Response (EIDSR) platform for outbreak monitoring, and Yobe deployed NAVISION and M-Supply for logistics and financial management. These systems were often implemented with donor support and operated in silos, with little or no integration across platforms.

Most states’ facility and patient registries were present but were often incomplete or poorly maintained. Many states reported being part of the National Health Facility Registry (HFR), but GIS coordinates were frequently missing or obsolete. Some states, like Borno and Yobe, had undertaken geo-tagging of facilities through external support, yet the registries were often not linked to broader systems or routinely used. The availability of individual digital identities for patients was almost non-existent. Respondents noted that while some platforms collected demographic data, secure and unified patient identifiers, such as Master Patient Indexes, were not established.

### 3.3 State ranking across the Maturity spectrum

As shown in Figure 3 below, only Lagos state was in the early adoption stage of digital health maturity. This was driven by relatively strong performance in services and applications, political commitment, and early investments in interoperability through the Smart Health Information Platform (SHIP). Notably, the maturity scoring for Lagos was informed by stakeholder interviews with donor and private sector partners due to the unavailability of government officials at the time of data collection. While this triangulated approach allowed for sufficient evidence capture, it introduces a minor limitation in comparability with other states.

**Figure 3:**
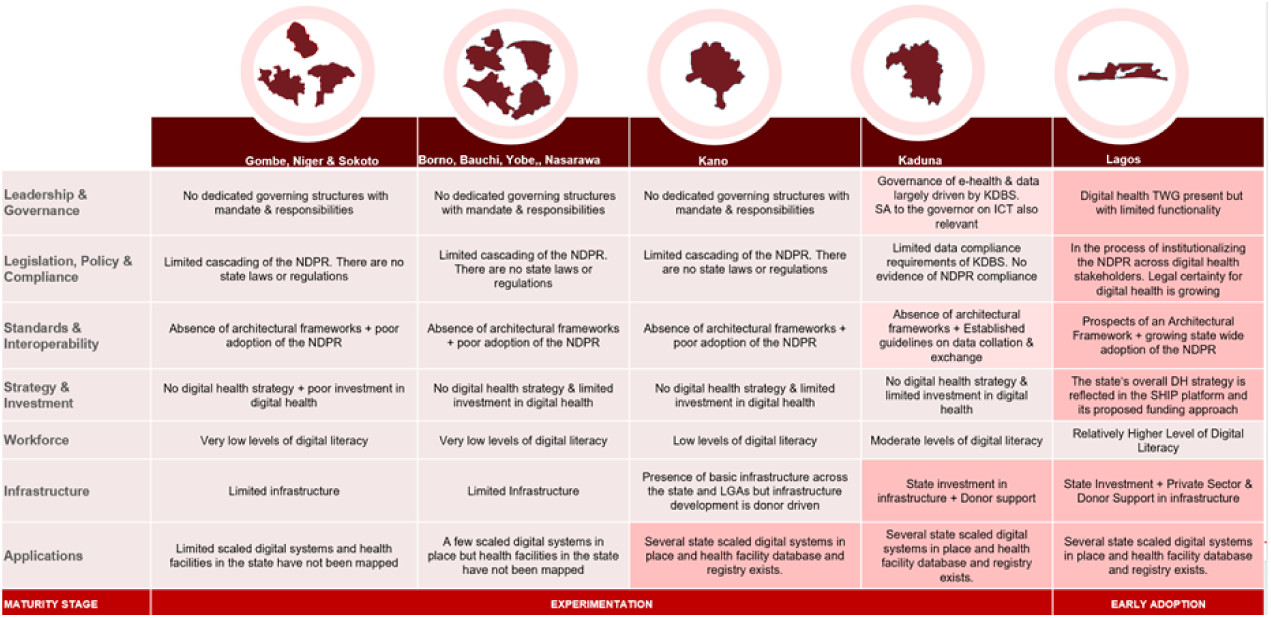
Digital health maturity classifications across the 10 states

Kaduna and Kano States followed, both classified within the late phases of the experimentation stage. These states showed more substantial traction in deploying applications, infrastructure support, and private sector engagement, although governance and legislative structures were still evolving. In Kaduna, platforms like the Health Facility Analysis (HEFA) and the data quality assurance (DQA) dashboards were actively used. At the same time, Kano demonstrated some investment in digital platforms and data tools, albeit with less consistency.

**Figure 4:**
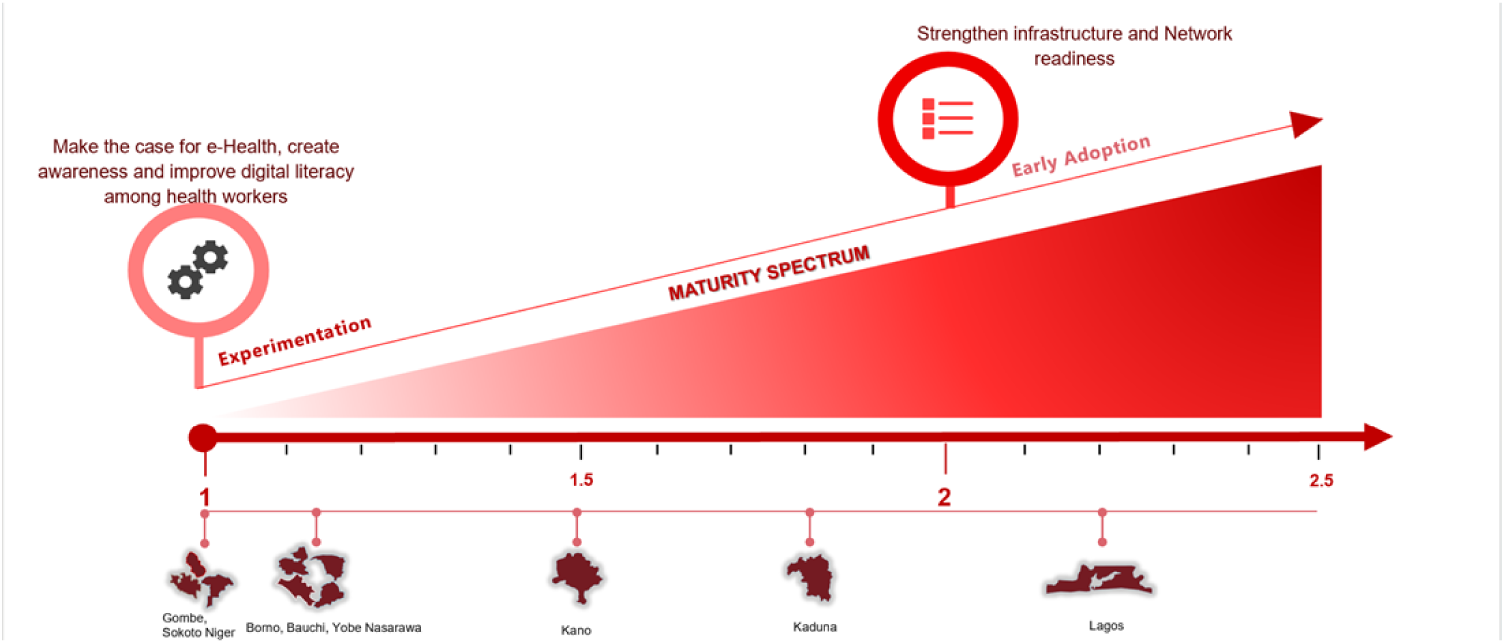
Positioning of States along the Maturity Spectrum

The remaining seven states, Borno, Bauchi, Gombe, Niger, Nasarawa, Sokoto, and Yobe, were all classified in the experimentation stage. This classification reflected minimal coordination, low investment, poor infrastructure, and reliance on donor-driven implementations. These states often lacked digital health strategies, institutional governance structures, or consistent training for the health workforce.

This spread reflects a broader pattern observed during the assessment—states with stronger enabling environments (Leadership/governance, legislation, strategy, investment, and standards/interoperability) tended to be more mature, while those with weaker governance arrangements and limited investment remained in the early phases of digital health maturity.

### 3.4 Thematic Findings from the Profiling of Digital Health Interventions

**Table 3:**
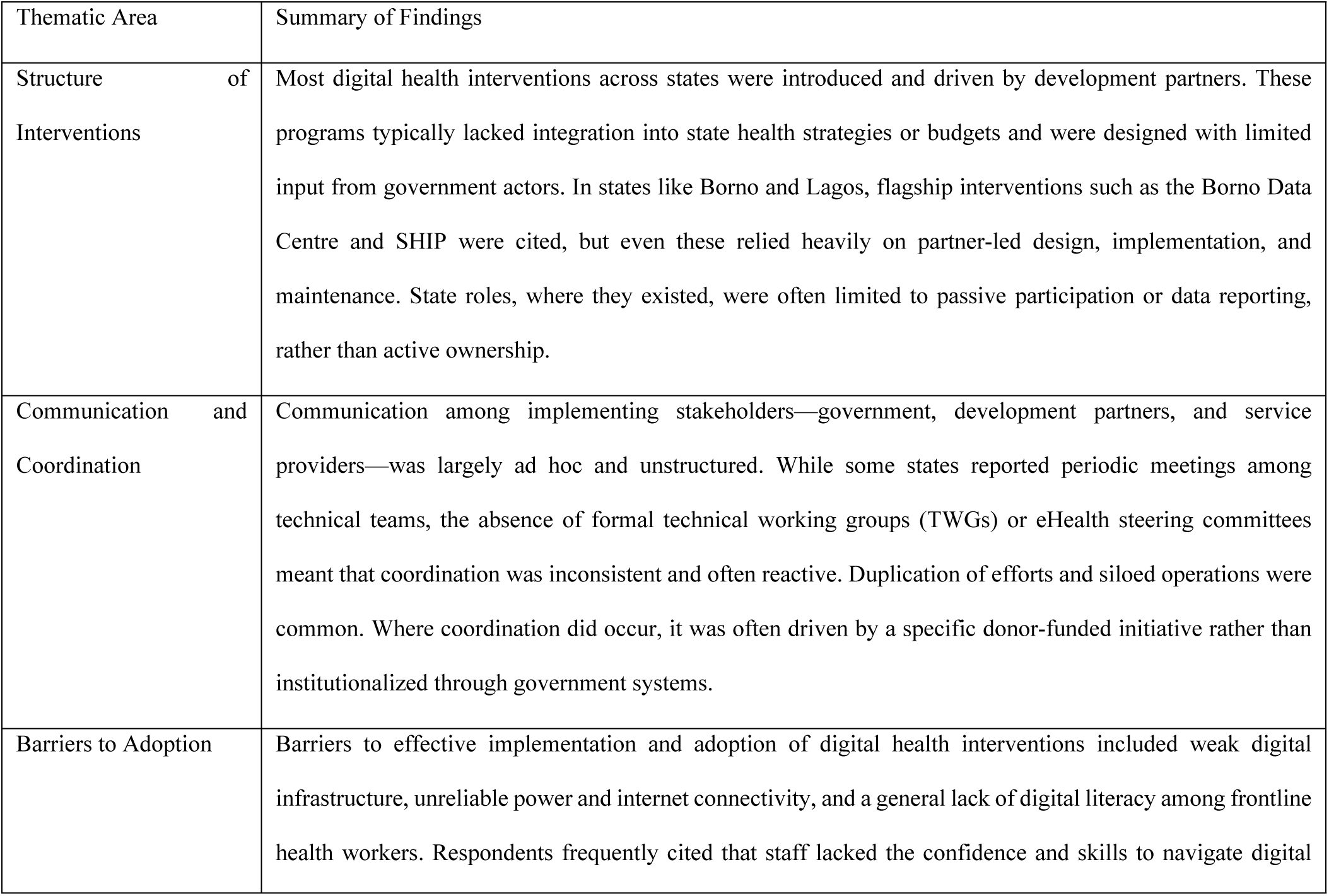
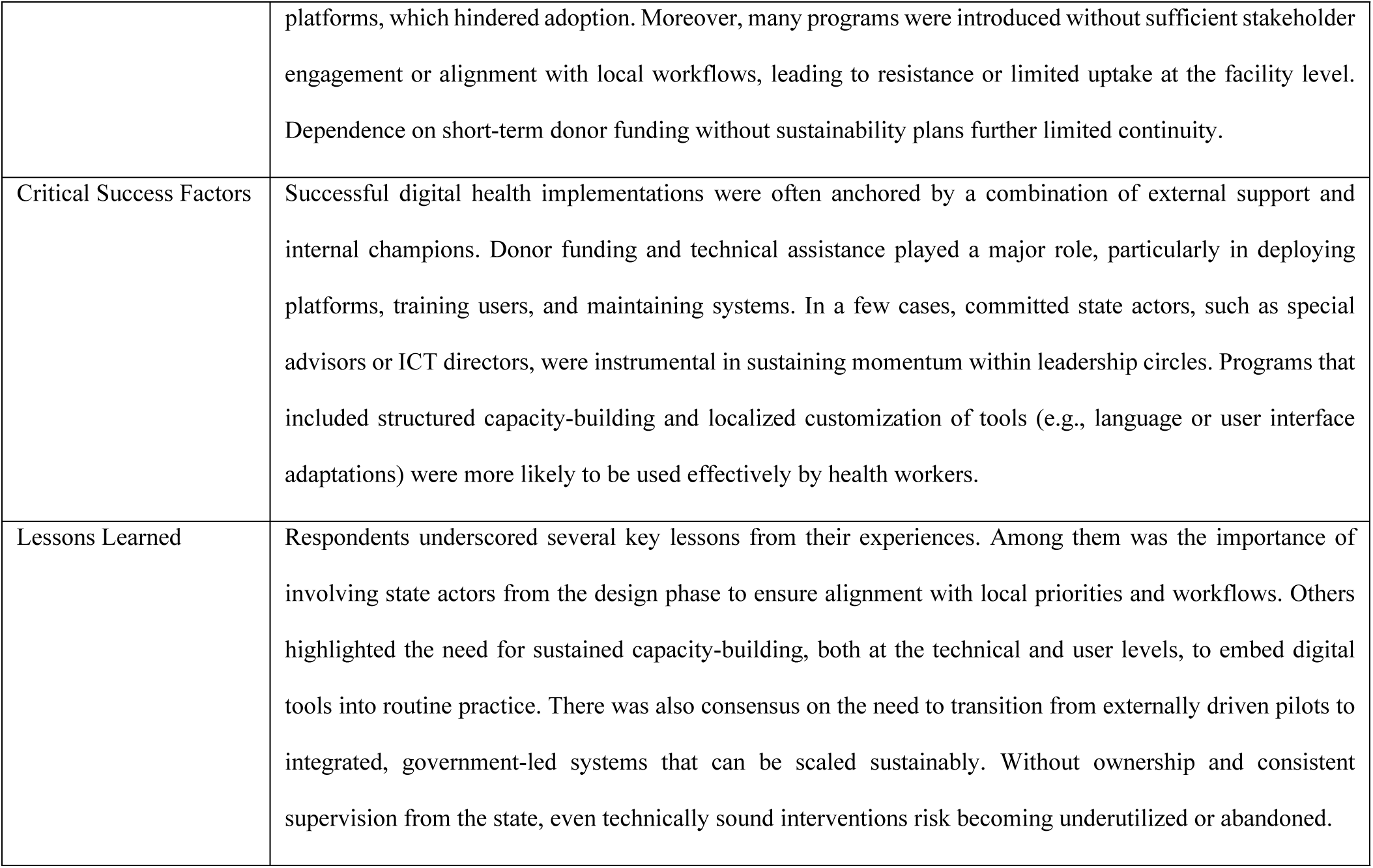
Thematic Findings from Part B – Digital Health Interventions.

## 4.0 Discussion

This paper provides a foundational look into the digital health maturity of ten Nigerian states, using the Global Digital Health Index (GDHI) with UN Foundation/ICT4SOML frameworks as a lens to analyse digital health maturity. Although the assessment was conducted in 2021, its insights remain timely, serving as a baseline for current reforms and as a mirror for how far state-level digital health ecosystems have evolved in 2025. The analysis underscores key issues related to governance, investment, infrastructure, digital literacy, and interoperability—all of which are central to Nigeria’s evolving digital health transformation agenda.

### Aligning with the GDHI Framework: A Pathway to Maturity

The use of the GDHI and UN Foundation/ICT4SOML frameworks was more than a methodological choice—it offers a structured, globally recognized pathway for digital health maturity at the subnational level. Alignment with GDHI principles encourages states to go beyond ad hoc, project-based deployments and instead build resilient systems based on coordinated leadership, policy coherence, sustainable financing, and integrated service delivery-critical elements of a strong enabling and ICT environment. The seven GDHI domains reflect not only technical readiness but also the institutional maturity required to sustain digital transformation. Positioning states along a maturity spectrum helps provide a roadmap for policy actions to drive progressive digital health reforms.

For Nigerian states, formal adoption of GDHI-aligned assessments could support internal benchmarking, drive local accountability, and feed into national discussions under the Nigeria Digital in Health Initiative (DIHI). This is particularly important as states are at different levels of digital health maturity.

### From 2021 to 2025: Contextual Continuity and Change

While four years have passed since the data were collected, the state of digital health in Nigeria continues to reflect many of the dynamics observed in the assessment. Fragmentation, limited institutional ownership, and donor-dependency remain key challenges in many states. However, the groundwork laid by early assessments like this one has helped shape the broader policy dialogue and prioritize state-level capacity strengthening.

Since 2021, Nigeria has made visible progress in digital health, marked most notably by the digital health strategy (2021-2025) and the recent Nigeria Digital in Health Initiative (NDHI), which places standards and interoperability at the core of national reform. The NDHI is a federal government-led effort to transform Nigeria’s healthcare system through digital technology, aiming to improve healthcare accessibility, efficiency, and quality nationwide^14,15^. At its core, it seeks to implement a national digital health architecture with interoperable electronic medical records (EMRs).

This is a critical shift, as many states assessed in this study lacked any digital health architecture or functional data exchange protocols. NDHI now promises an opportunity to resolve this fragmentation, especially if state governments are empowered to align their systems with national interoperability guidelines.

### Solving from first principles: Classifying digital systems using a context-sensitive taxonomy that enables interoperability

The challenges revealed in this assessment—particularly the fragmentation of digital platforms, the absence of governance mechanisms, and limited interoperability—highlight the urgent need for a structured, system-wide approach to digital health. The Nigeria Digital in Health Initiative (NDHI) offers an important national framework to guide this transition, especially around establishing standards, enforcing interoperability, and formalizing governance.

However, for states to operationalize NDHI effectively, they will first require a practical and context-sensitive framework to guide how they classify or define digital health. The Gates Foundation’s working taxonomy for digital health at scale provides a useful blueprint^16^. It was developed based on the need to identify with the systems at the most foundational level, that is, supporting infrastructures that drive digital health initiatives, while also addressing use cases surrounding health.^17^ As shown in Figure 5 below, the taxonomy maps out digital health across three layers: digital use cases (prevention, case management, surveillance), digital infrastructure (including identifiers, EHRs, registries, HR systems), and the foundational enablers (connectivity, identity, payments, cybersecurity).

**Figure 5:**
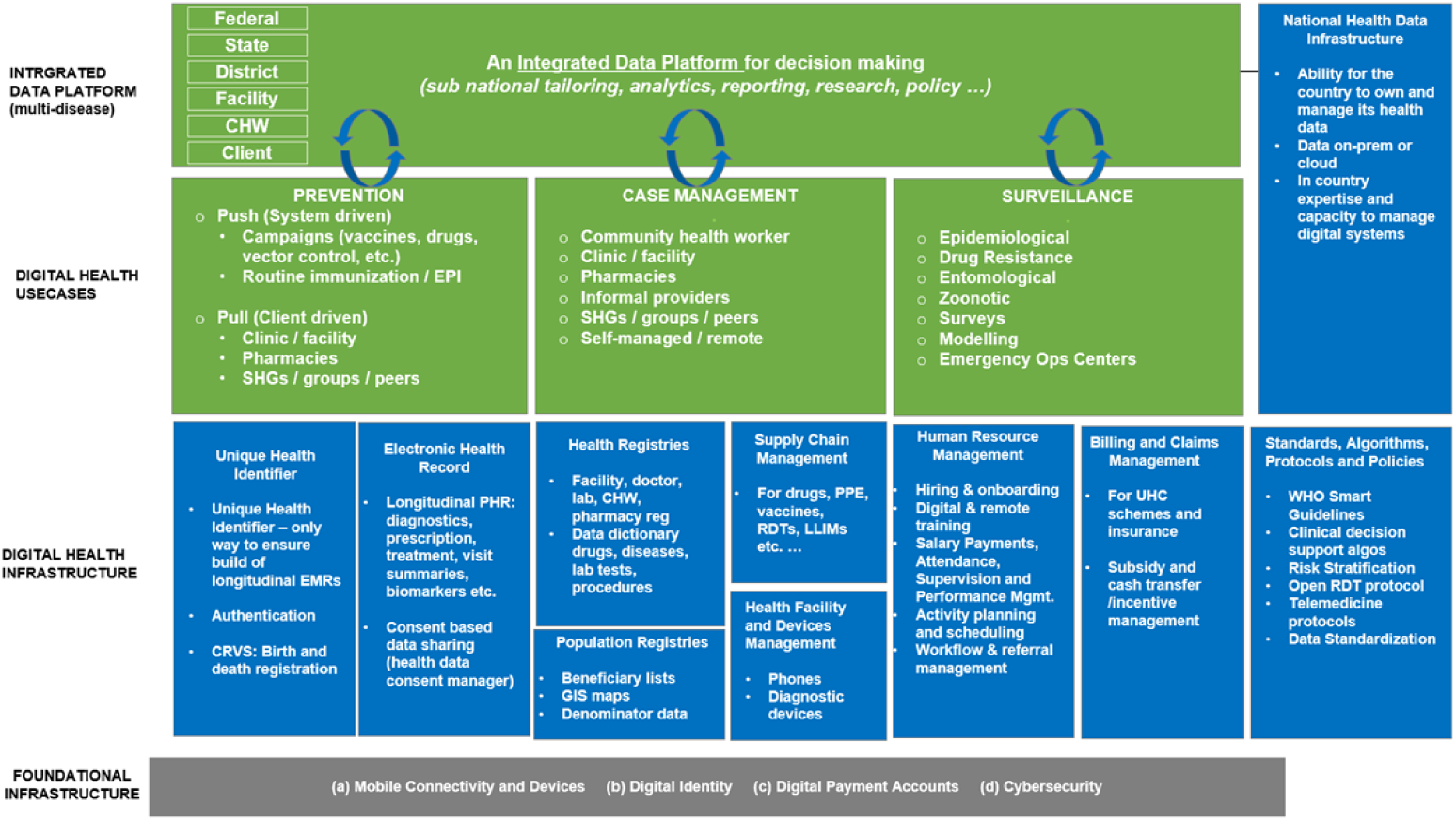
The Gates Foundation Working Taxonomy

This structured framing is particularly relevant for Nigeria’s federated health system, where digital health solutions must function across multiple levels—from federal to local government service points and down to individual frontline health workers. For instance:

- Integrated data platforms should be designed with sub-national tailoring in mind, enabling states to use analytics for localized decision-making and reporting.
- Digital health infrastructure—such as unique health identifiers, GIS-enabled registries, interoperable electronic medical records proposed by the NDHI—can support continuity of care and resource tracking.
- Use cases in prevention, case management, and surveillance need to be mapped explicitly to state health priorities and capabilities and supported with scalable platforms.

By aligning their planning and investment with this taxonomy, states can move beyond donor-led pilots and fragmented tools to institutionalized, scalable digital ecosystems.

### Digital Literacy: Bridging the Human Infrastructure Challenge

The sub-national assessment of digital health maturity in Nigeria reveals a pervasive gap in digital literacy among health workers, significantly undermining the effectiveness of digital health interventions. Across the ten states assessed, it was evident that digital tools, even when available, were underutilized due to limited ICT competencies among frontline health workers and program staff. This gap is not merely a skills deficit—it represents a structural barrier to digital health scale-up, at the same level as infrastructure or connectivity challenges. The findings indicate that digital health capacity-building initiatives have mainly been fragmented, focusing predominantly on program-specific tools like DHIS2 or vertical surveillance platforms, with minimal emphasis on holistic ICT skills or digital fluency.

The absence of structured pre-service training in digital health across most states compounds the challenge. Health professionals typically enter the workforce without exposure to health information systems. In-service training, where it exists, is infrequent and targeted, leaving significant gaps in the workforce’s ability to engage with digital platforms effectively. For instance, in states like Gombe, Nasarawa, and Yobe, where workforce readiness scored the lowest, digital health interventions struggled to gain traction due to health workers’ unfamiliarity with basic ICT operations.

Addressing this literacy gap requires more than isolated, donor-led training programs; it calls for integrating digital health and digital literacy into pre-service education alongside structured in-service mentorship and digital job aids. Pre-service training should be standardized to include core competencies in health information systems, digital record management, and telemedicine applications. Concurrently, in-service training should evolve into structured mentorship programs, reinforced by digital job aids and continuous learning opportunities. This reimagining of digital literacy as foundational "human infrastructure" is critical for sustainable digital health implementation, in the Nigerian context, the Digital in Health Initiative. Without robust digital skills among health workers, even the most advanced digital health platforms risk underutilization, reducing their potential impact on service delivery and health outcomes.

### Digital Infrastructure remains a critical but misunderstood barrier

This landscape assessment revealed a critical misunderstanding among respondents across surveyed states regarding the infrastructure necessary for digital health systems. Findings indicate that respondents often misconstrue foundational infrastructure for digital health infrastructure. This misinterpretation was evident in the focus on power supply, internet connectivity, hardware, and maintenance as key infrastructural challenges. While these are indeed vital, they represent only the foundational layer necessary to support digital health.

In Figure 5, the Gates Foundation’s taxonomy^16^ classifies infrastructure into two distinct but interdependent categories: Foundational Infrastructure and Digital Health Infrastructure. Foundational infrastructure consists of core elements such as mobile connectivity, digital identity, digital payment accounts, and cybersecurity components that create the digital foundation for health technology to function. For instance, robust mobile networks are essential not just for telemedicine but for real-time data collection in surveillance and case management. Similarly, digital identities enable unique patient identifiers crucial for electronic health records (EHRs).

In contrast, digital health infrastructure builds upon these foundations to support health-specific applications and digital health use cases. This includes EHRs for clinical documentation, health registries for population data, supply chain management systems for medical commodities, and Human Resource Information Systems (HRIS) for health workforce planning and coordination.

The assessment findings, however, show that many respondents associated infrastructural challenges with visible, tangible issues such as hardware (computers, tablets) and power, etc. Foundational infrastructure, such as digital identity systems and cybersecurity, and digital health-specific infrastructure, such as EHR, health registries, and human resource management systems, were rarely mentioned. This gap in understanding also points to broader digital literacy issues where stakeholders focus on what they know, can physically see and interact with, rather than recognizing the deeper layers of infrastructure needed to make digital health systems truly effective.

### Political Will and Budgeting as Enablers

Political will remains an essential enabler for digital health success at the state level, serving as the foundation upon which planning, budgeting, and governance structures are built. The assessment findings reveal that states with stronger political commitment, such as Lagos and Kaduna, demonstrated more visible progress in digital health implementation. In Lagos, for instance, political support facilitated the development of the Smart Health Information Platform (SHIP), which is designed to serve as an interoperability layer for all health information systems in the state. In Kaduna, investments in digital health were reflected in strategic platforms such as the Health Facility Analytics (HEFA), even though these efforts were primarily donor driven.

However, political commitment alone is insufficient without integration into state annual operational plans (AOPs) and corresponding budgetary allocations. The assessment found that in many states—such as Bauchi, Borno, Gombe, Yobe, Niger and Nasarawa—digital health is not represented in annual budgets and states also lack costed digital health action plans, which hinders their capacity to mobilize internal resources and sustain digital health initiatives post-donor funding. In contrast, Lagos and Kaduna demonstrated some progress in allocating resources for ICT infrastructure to support digital health projects. Nevertheless, this allocation remains inconsistent across other states, with the majority relying on short-term, externally funded projects not embedded into the fiscal planning cycles.

We also see that functional digital health Technical Working Groups (TWGs) are notably absent in most states, except for Lagos, which has a structured TWG that meets regularly to coordinate digital health activities. In many other states, digital health leadership is informal or non-existent, limiting strategic oversight and accountability for execution. The absence of TWGs constrains states’ ability to coordinate digital health investments, harmonize donor contributions, and advocate for political prioritization at higher levels of government.

## 5.0 Study Limitations

This study was designed to provide a comprehensive assessment of digital health maturity across ten Nigerian states using a combination of document reviews and key informant interviews (KIIs). While the methodology was robust, several limitations should be noted.

First, the study encountered challenges accessing complete and up-to-date documentation at the state level. Many digital health initiatives lacked publicly available reports or formal evaluations, limiting the desk review’s scope and depth. In several states, documentation of past digital health investments was either unavailable or held informally by implementing partners. As a result, the study leaned more heavily on key informant interviews to supplement gaps in the secondary data.

Second, there were language and comprehension barriers during the KIIs, particularly in some states. While most interviews were conducted in English, nuances and comprehension may have been better captured had interviews been conducted in local languages such as Hausa and translated during transcription. This may have affected the consistency and depth of some responses.

Third, there were non-responses from key government stakeholders in Lagos State due to an ongoing internal e-health assessment, which the state gave greater priority to. To mitigate this, data for Lagos was triangulated using interviews with donor and private sector stakeholders who were actively implementing programs in the state. While this ensured that Lagos was not excluded from the analysis, it introduces a limitation in comparability, as other states’ assessments relied directly on government perspectives.

Additionally, respondents displayed varying levels of digital literacy, impacting the depth of discussions about certain digital health concepts. In some states, respondents equated digital health solely with tools like WhatsApp or the use of mobile phones and computers, lacking awareness of broader systems, governance structures, or interoperability standards. This limited the granularity of insights, especially in areas like legislation, standards, and architecture.

Finally, the subjectivity of the scoring process, while mitigated by a structured abstraction and validation method, remains a consideration. In domains where multiple respondents provided differing inputs, the scoring relied on the mean scores based on the responses. More weight was given to documented evidence over personal recall to reduce bias, but the potential for residual subjectivity cannot be entirely eliminated.

Despite these limitations, the study presents one of Nigeria’s most comprehensive subnational digital health maturity assessments to date. It offers valuable insights for policy, investment, and future research.

## 6.0 Conclusion and Recommendations for Advancing Digital Health Maturity in Nigeria

This assessment has shown that the ten (10) states are at different stages of digital health maturity. While some pilot foundational systems, others invest in platforms and explore interoperability at scale. These varying readiness levels underscore the need for differentiated yet coordinated actions. The following recommendations are designed to accelerate progress across the maturity spectrum and strengthen digital health ecosystems across the surveyed states and nationwide.

### I. Institutionalize Digital Health Governance Mechanisms at the State level

There is an imperative for states to establish, formalize, or revitalize digital health Technical Working Groups (TWGs) domiciled within the Ministry of Health, to coordinate actors, streamline planning, and ensure that digital interventions align with state priorities. These TWGs should be multi-sectoral, include private sector and civil society representation, and be integrated into the state’s broader health governance architecture. The TWGs can serve as governance hubs facilitating proactive policy development, implementation oversight, and stakeholder coordination. The TWG’s activities should be integrated into the broader health governance architecture by aligning it with Annual Operational Plans (AOPs) and long-term state health strategies.

### II. Develop and implement context-specific digital health strategies aligned with the AOP processes

State Ministries of Health should be supported to develop digital health strategies aligned with their respective maturity levels, whether at the experimentation, early adoption, or developing, scaling up, and mainstreaming stage. These digital health strategies should be embedded within State Health Sector Strategic Plans and Annual Operational Plans (AOPs). National coordination should allow for differentiated implementation support based on each state’s trajectory, rather than a one-size-fits-all model.

In addition, states should move beyond ad hoc funding of ICT to institutionalized digital health budgeting. This will involve collaboration with the Ministry of Budget and Planning and the Ministry of Finance to create and execute on clear and dedicated budget lines for digital health infrastructure, tools, capacity building, and system maintenance. This structured financial planning will enhance transparency, sustainability, and enable better tracking of return on digital investments.

### III. Strengthen Legal and Regulatory Foundations for Digital Health

To drive compliance and scale digital health interventions, States must adapt and domesticate national digital health policies, such as the National Digital Health Strategy (NDHS) and the Nigeria Digital Health Initiative (NDHI). This domestication process should be guided by the appropriate legislative, executive, or administrative mechanisms, driven by the state-level digital health TWGs, whose activities towards adapting or adopting national strategies should be planned for and budgeted. In addition, building awareness and compliance capacity among public officials and health workforce is also essential to ensure adherence to the Nigeria Data Protection Regulation (NDPR). Additionally, states should establish enabling legal environments that support innovation, including using regulatory sandboxes to safely and efficiently pilot emerging digital health solutions.

### IV. Advance Standards and Interoperability through Structured System Building

To achieve resilient and interoperable digital health ecosystems, states should align with national guidelines under the Nigeria Digital in Health Initiative (DIHI) while leveraging structured implementation frameworks such as the Gates Foundation’s digital health taxonomy. This dual alignment will help organize state-level ecosystems around three core layers: use cases (e.g., surveillance, case management, vaccine logistics etc), digital infrastructure (e.g., registries, ID systems, health data warehouses), and foundational enablers (e.g., broadband connectivity, cybersecurity).

States should be supported to build, adapt, or align with standards-compliant digital health architectures that allow seamless integration across platforms, facilities, and programs. Digital health initiatives such as the NDHI must prioritize data harmonization—including standardized terminologies, APIs, and interoperability protocols—while ensuring linkage to nationally scaled platforms such as the National Health Management Information Systems (NHMIS). National standards should be translated into practical, context-sensitive guidance for states to adapt or adopt without undermining local innovation or ownership. By doing so, Nigeria can create a more connected, scalable, and sustainable digital health system across all tiers of government.

### V. Integrate Digital Health into Health Workforce Development

Human capacity is central to digital health execution. States must begin to cultivate and prepare their human infrastructure for digital health. First, digital health competencies must be embedded into pre-service and in-service training curricula for health workers across cadres. Curricula should include structured digital literacy programs that cover all dimensions of digital literacy: technical skills, information literacy, communication literacy, data literacy, media literacy, security literacy, digital problem-solving, critical thinking, and cultural understanding. The Federal Ministry of health can collaborate with the Federal Ministry of Education and ICT, training institutions, and digital health startups to revise health curricula and incorporate digital modules around a core set of nationally scaled digital health platforms such as DHIS-2, SORMAS(Surveillance, Outbreak Response Management and Analysis System), etc Secondly, digital health career pathways within the public service should be clearly defined, with established roles, job descriptions, and performance incentives. Furthermore, government and partners should scale targeted digital literacy initiatives for frontline workers, M&E staff, and program managers as part of in-service training to ensure system uptake and quality data use. This can be done by integrating digital courses and digital learning into Continuous Professional Development (CPD) mechanisms so that digital skills are continually updated and applied.

### VI. Test bundled infrastructure solutions through innovator states

In line with the NDHI implementation, the Nigerian government, in collaboration with partners, may pilot bundled infrastructure solutions to address key gaps in power, connectivity, and hardware across a few innovator states such as Lagos, Kaduna, FCT, and Rivers. This is in keeping with an incremental implementation of NDHI using potential innovator states based on Rogers’ innovation diffusion curve^18^. This strategy may encompass:

`O` Expanding NPHCDA’s ongoing solarization efforts across Primary Healthcare Centres (PHCs) to Secondary health facilities ensures stable and sustainable power supply in health facilities in pilot states.
`O` Partnering with NIGCOMSAT and major telecom providers to deploy broadband access in pilot states, focusing on last-mile connectivity to improve data capture especially in remote facilities.
`O` Pursuing hosting and cloud solution collaborations with Galaxy backbone for hosting, storage and evolutive maintenance of digital health infrastructure.
`O` Exploring partnerships with private sector hardware providers and donor partners to address hardware deficits, ensuring that facilities are equipped with the necessary devices for digital health applications.

By leveraging these innovator states as proof-of-concept sites, Nigeria can validate scalable digital health infrastructure models that can be replicated in other regions, accelerating the path to nationwide digital health maturity.

### VII. Maximizing digital health impact using nationally scaled digital solutions

State governments and their partners must realize that to maximize the impact of digital health on service delivery, states must prioritize and scale up solutions that align with their digital health trajectory and address priority digital health use cases. A critical consideration for sustainability is ensuring that all digital applications are designed with state relevance, interoperability, and long-term integration with the national system in mind. Too often, states and their development partners implement standalone platforms which are poorly aligned with national data systems, resulting in duplication, fragmentation, and inefficiencies. In line with priority 12 of the Sector Wide Approach for Health (SWAp) pillars^5^, all deployed tools—whether donor-funded or locally developed—must conform to national standards and be compatible with overarching platforms such as the National Health Management Information System (NHMIS).

The District Health Information Software 2 (DHIS2)—which powers Nigeria’s NHMIS—remains the most scaled and standardized digital health platform in the country. It offers a suite of use cases and modules (some activated, others not yet implemented in Nigeria) that can address many of the functionalities that states often seek to build independently. To prevent the proliferation of siloed systems, strengthen health data collection, and reduce the risk of states investing in duplicative platforms, the FMOHSW and the SWAP Coordination office and its partners must strengthen the management of DHIS2 at the national level, with decentralized access at the state level. This includes creating a dedicated budget line, sustaining a national technical support team, and ensuring that states have adequate access, training, and autonomy to manage their relevant modules. Doing so would preserve system coherence and ensure national investments are fully leveraged at the subnational level.

### VIII. Strengthen Local Innovation and Public-Private Partnerships

States can create enabling environments for local innovation and public-private partnerships (PPPs). By actively engaging startups, ICT hubs, research institutions, and health technology firms, states can co-create tools that are contextually appropriate, user-centered, and responsive to service delivery gaps. States such as Lagos offer early proof-of-concept that with supportive governance, aligned incentives, and a vibrant innovation ecosystem, digital tools can be deployed and scaled effectively. However, to truly advance on the digital maturity spectrum, states must move beyond ad-hoc implementation to developing concrete enablers for innovation. This includes establishing innovation-friendly procurement processes, providing institutional clarity on collaboration mechanisms, and adopting co-investment models that reduce risk for startups addressing public health issues using technology. These steps will stimulate local digital health ecosystems and help close persistent digital health gaps that the government may not address quickly.

### IX. Establish a National Mechanism for State Benchmarking and Peer Learning

To sustain momentum in digital health advancements, a National Digital Health Observatory should be established within the FMOHSW to track digital health maturity and monitor implementation progress across all 36 states and the Federal Capital Territory (FCT). This mechanism would enable real-time visibility into state-level achievements, gaps, and trajectories. Furthermore, it would be the foundation for structured peer learning networks, where states can share best practices, tools, and strategies. Such collaborative platforms would be particularly valuable for lower-maturity states, allowing them to leapfrog common barriers through access to tested solutions and targeted support.

As a critical next step, a comprehensive digital health landscape study should be conducted across the 36+1 states to establish a robust baseline for benchmarking. This study would provide a clear and recent understanding of the digital health ecosystem across the country, facilitating the development of targeted interventions and enabling progress tracking over time. The findings from the study would inform the implementation, collaboration, and foster a culture of accountability and continuous improvement in digital health initiatives nationwide.

## 7.0 Declarations

- **Funding**: This work was funded by the Gates Foundation.
- **Conflicts of Interest**: The authors declare no conflicts of interest.
- **Ethical Approval**: Approval was obtained from National Health Research Ethics Committees.
- **Consent**: Informed written consent was obtained from all participants.
- **Author Contributions:**
  - Chukwuemeka Azubuike *(Devstork Platform for Development)* led the technical delivery of the study, including conceptualization, stakeholder engagement, oversight of data collection, data analysis, and development of the initial manuscript draft. He also integrated feedback from co-authors and reviewers into revised versions of the manuscript.
  - Ramat Seghosime *(Devstork Platform for Development)* contributed to data interpretation and supported refinement of the manuscript, with a focus on analytical rigour and clarity of presentation.
  - Chike Nwangwu *(NOI Polls)* oversaw project commissioning and provided strategic input during the design and validation phases. He also contributed to manuscript review and quality assurance, ensuring alignment with funder expectations.
  - Precious Nwiko *(Independent Digital Health Expert)* provided a second-level expert review of the findings and manuscript, validating the digital health framing and enhancing the analytical depth of the discussion and recommendations.
  All authors reviewed and approved the final version of the manuscript.
- **AI Use Disclosure**: The authors affirm that this manuscript was conceived, analysed, and written by the authors. Artificial intelligence tools (specifically [e.g., ChatGPT, Perplexity) were used solely for language refinement, grammar correction, and sentence formatting suggestions during the drafting process. No AI tools were used to generate or analyze data, and all intellectual contributions—including conceptual framing and writing decisions—remain the authors’ sole responsibility.

## 8.0 Supplementary Material: Scoring System

### Leadership & Governance and Strategy & Investment

#### Indicator 1: Digital health prioritized through governance

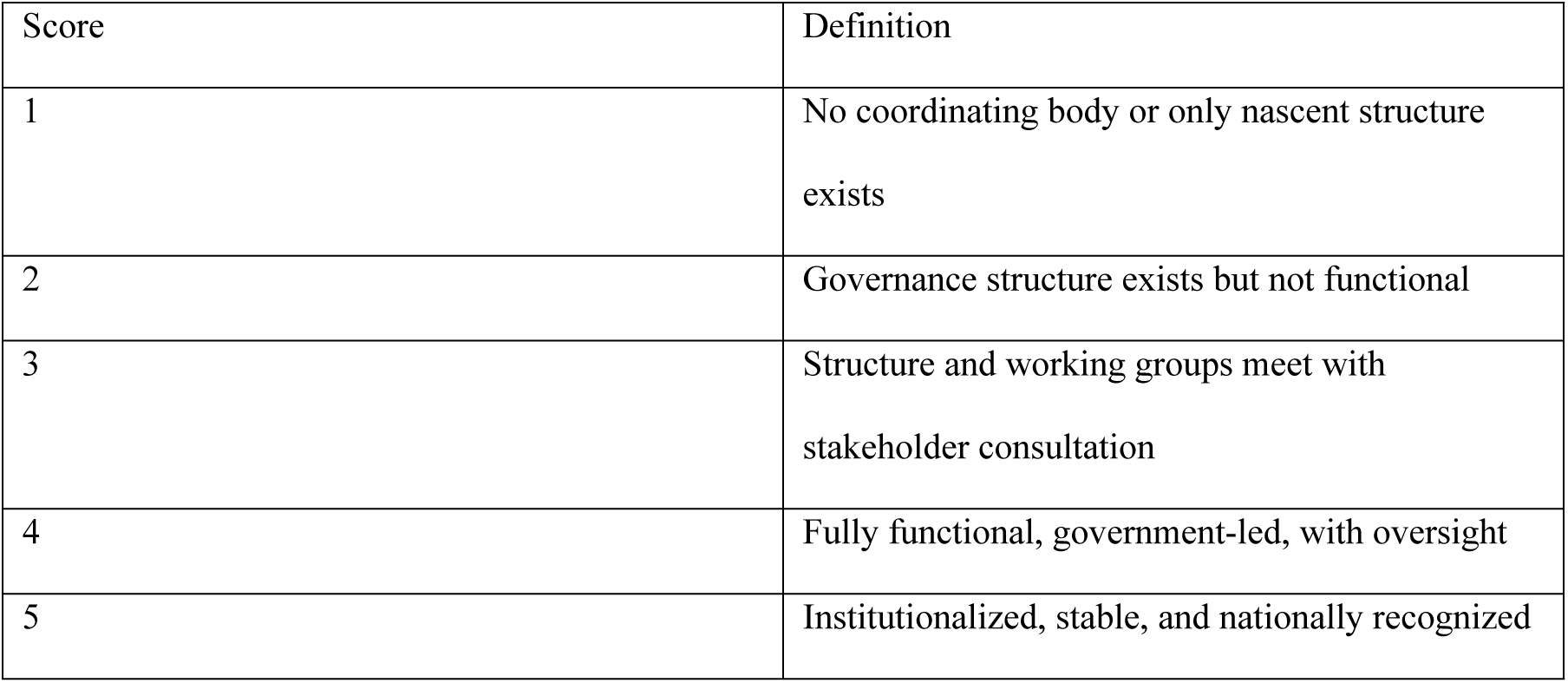

#### Indicator 2: Digital health prioritized through planning

| Score | Definition |
| --- | --- |
| 1 | Not included in strategy |
| 2 | Discussed or proposed |
| 3 | Included in strategy/plan |
| 4 | Being implemented in strategy |
| 5 | Periodically evaluated and optimized |

#### Indicator 3: National eHealth/Digital Health Strategy or Framework

| Score | Definition |
| --- | --- |
| 1 | No strategy |
| 2 | Strategy approved |
| 3 | Strategy and costed plan approved |
| 4 | Strategy and plan partially implemented |
| 5 | Fully implemented and forward planning underway |

#### Indicator 4: Public funding for digital health

| Score | Definition |
| --- | --- |
| 1 | No budget line |
| 2 | <1% |
| 3 | 1–3% |
| 4 | 3–5% |
| 5 | >5% |

### Legislation, Policy & Compliance

#### Indicator 5: Legal Framework for Data Protection

Legislation, Policy & Compliance Indicator 5: Legal Framework for Data Protection
| Score | Definition |
| --- | --- |
| 1 | No law exists |
| 2 | Law proposed and under review |
| 3 | Law passed but not implemented |
| 4 | Law implemented but not consistently enforced |
| 5 | Law enforced consistently |

#### Indicator 6: Laws for privacy, confidentiality and data access

| Score | Definition |
| --- | --- |
| 1 | No law |
| 2 | Law proposed and under review |
| 3 | Law passed but not yet implemented |
| 4 | Law implemented but not enforced |
| 5 | Law enforced consistently |

#### Indicator 7: Protocol for regulating digital health services

| Score | Definition |
| --- | --- |
| 1 | No protocols or frameworks |
| 2 | Protocols/frameworks proposed |
| 3 | Protocols/frameworks passed but not fully implemented |
| 4 | Implemented but not consistently enforced |
| 5 | Implemented and enforced consistently |

#### Indicator 8: Cross-border data sharing security

| Score | Definition |
| --- | --- |
| 1 | No protocols or frameworks |
| 2 | Proposed but under review |
| 3 | Passed but not fully implemented |
| 4 | Implemented but not enforced |
| 5 | Enforced consistently |

### Capacity Building/Workforce

#### Indicator 9: Digital health in pre-service training

Capacity Building/Workforce Indicator 9: Digital health in pre-service training
| Score | Definition |
| --- | --- |
| 1 | No integration in pre-service training |
| 2 | Integration proposed |
| 3 | Approved but not implemented |
| 4 | Implemented but not enforced |
| 5 | Implemented and enforced |

#### Indicator 10: Digital health in in-service training

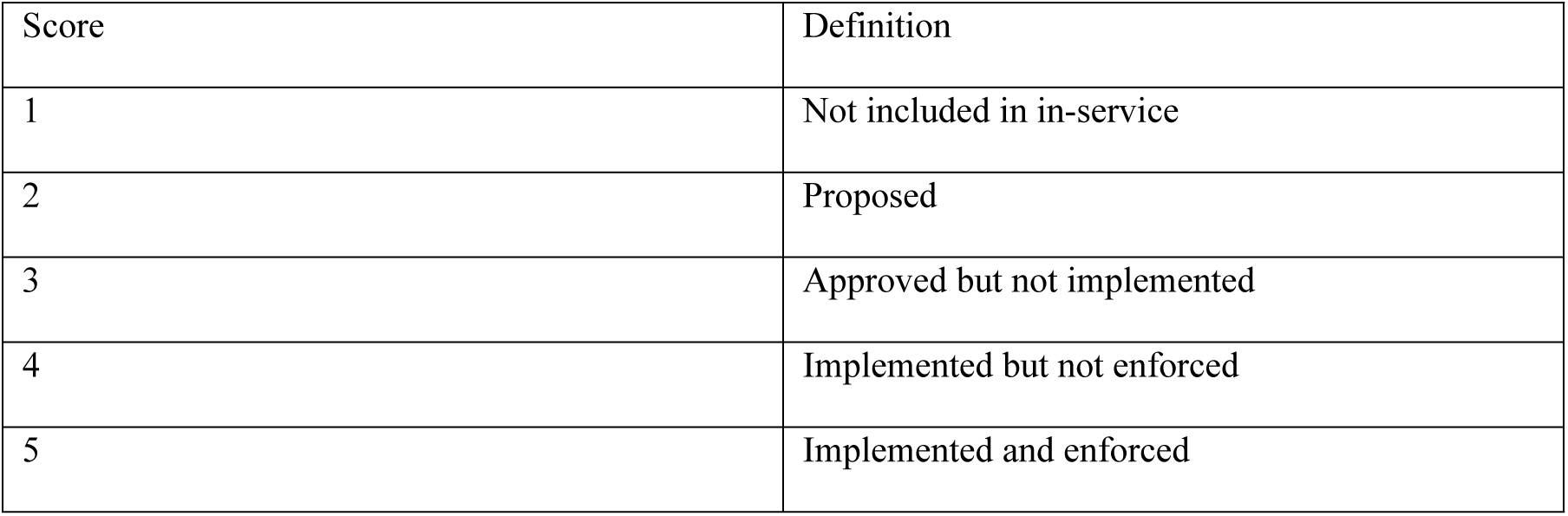

| Score | Definition |
| --- | --- |
| 1 | Not included in in-service |
| 2 | Proposed |
| 3 | Approved but not implemented |
| 4 | Implemented but not enforced |
| 5 | Implemented and enforced |

#### Indicator 11: Training of digital health workforce

| Score | Definition |
| --- | --- |
| 1 | No formal training framework |
| 2 | Framework proposed |
| 3 | Framework approved but not implemented |
| 4 | Training implemented but inconsistently |
| 5 | Training implemented and enforced consistently |

### Standards & Interoperability and Infrastructure

#### Indicator 13: eHealth architecture or health info exchange

Standards & Interoperability and Infrastructure Indicator 13: eHealth architecture or health info exchange
| Score | Definition |
| --- | --- |
| 1 | No architecture or exchange exists |
| 2 | Proposed but not approved |
| 3 | Operable with core functions |
| 4 | Implemented with government leadership |
| 5 | Continuously updated and used in national strategy |

#### Indicator 14: Health information standards

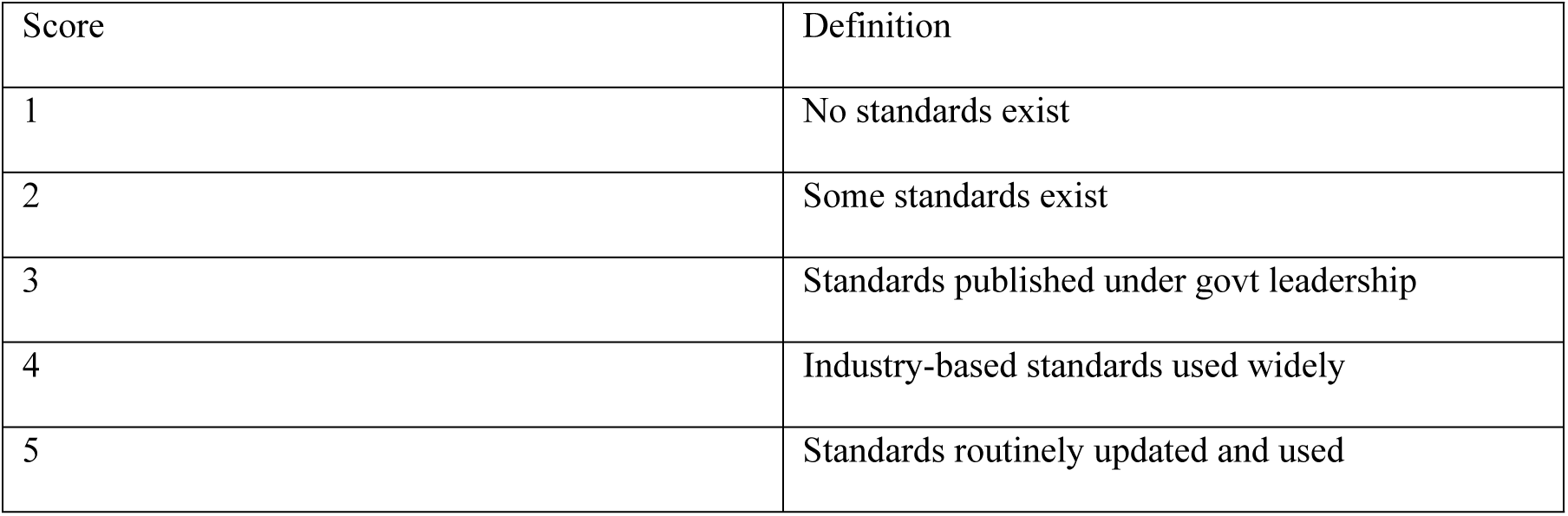

| Score | Definition |
| --- | --- |
| 1 | No standards exist |
| 2 | Some standards exist |
| 3 | Standards published under govt leadership |
| 4 | Industry-based standards used widely |
| 5 | Standards routinely updated and used |

#### Indicator 15: Network readiness

| Score | Definition |
| --- | --- |
| 1 | No infrastructure plan |
| 2 | Plan exists but not implemented |
| 3 | Plan implemented with minimal coverage |
| 4 | Plan implemented partially and consistently |
| 5 | Widely available and regularly upgraded |

#### Indicator 16: Support for infrastructure maintenance

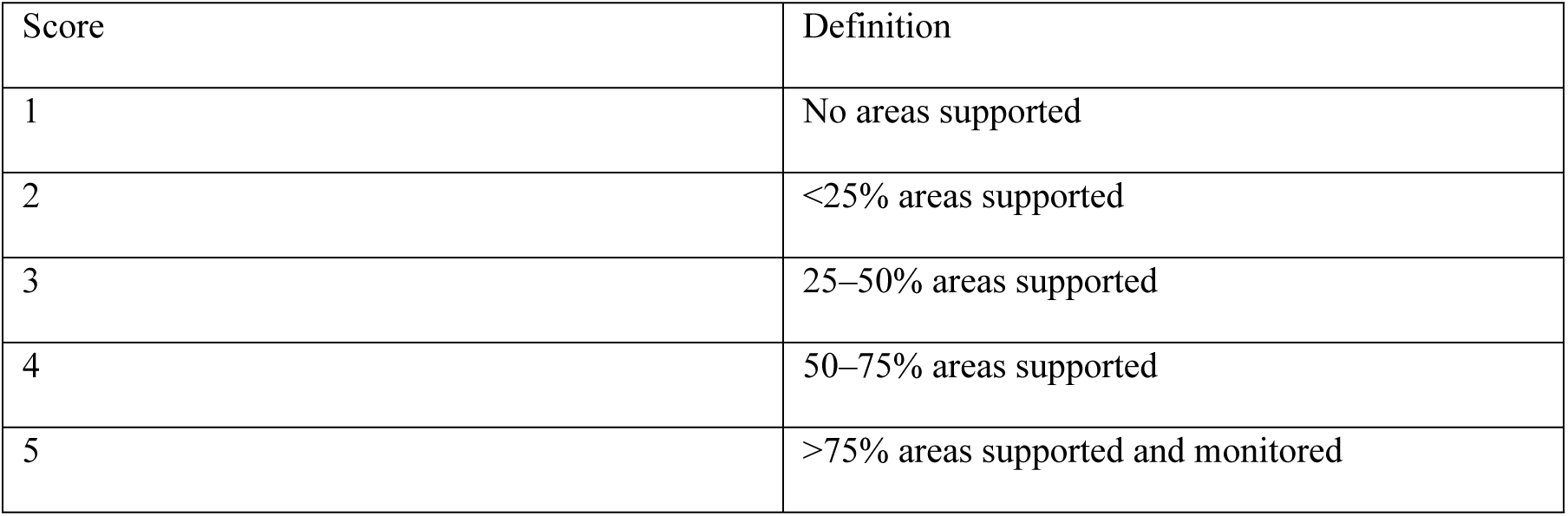

### Applications

#### Indicator 17: Nationally scaled digital health systems

Applications Indicator 17: Nationally scaled digital health systems
| Score | Definition |
| --- | --- |
| 1 | No priority areas supported |
| 2 | <25% areas supported |
| 3 | 25–50% areas supported |
| 4 | 50–75% areas supported |
| 5 | >75% areas supported and monitored |

#### Indicator 18: Digital identity and GIS mapping

| Score | Definition |
| --- | --- |
| 1 | Registries not available or current |
| 2 | Being developed but not usable |
| 3 | Partially available and used irregularly |
| 4 | Available and updated with geo-tagging |
| 5 | Routinely updated and used for planning |

## Data Availability

All data supporting the findings described in this manuscript are available in the article, Supplementary Information, or from the corresponding author upon request

## Footnotes

4 2015 was important as it aligned with the timeline of the National e-health strategy and also to ensure that collected data still retained its currency in the study year-2021.

5 Strengthen health data collection, reporting and usage – starting with the core indicators

## Notes

### Competing Interest Statement

The authors have declared no competing interest.

### Funding Statement

Yes

### Author Declarations

Ethical approval was obtained from the National Health Research Ethics Committee at the Federal Ministry of Health (FMOH).

